# Infant Subcortical Brain Volumes Associated with Maternal Obesity and Diabetes: A Large Multicohort Study

**DOI:** 10.1101/2025.03.25.25324641

**Authors:** Ann M. Alex, Jerod M. Rasmussen, Jetro J. Tuulari, Julie Nihouarn Sigurðardottir, Claudia Buss, Kirsten A. Donald, A David Edwards, Sonja Entringer, John H. Gilmore, Nynke A. Groenewold, Hasse Karlsson, Linnea Karlsson, Katherine E. Lawrence, Inka Mattilla, Dan J. Stein, Martin Styner, Paul M. Thompson, Pathik D. Wadhwa, Heather J. Zar, Xi Zhu, Gustavo de los Campos, Rebecca C. Knickmeyer, Shan Luo, the ENIGMA-ORIGINs working group

**Author notes:** Correspondence and reprint requests can be made to Dr. Shan Luo, PhD, Assistant Professor of Medicine, USC Keck School of Medicine, Division of Endocrinology and Diabetes, Diabetes and Obesity Research Institute, 2250 Alcazar Street; CSC 215, Los Angeles, CA 90089.

## Abstract

**Importance:** Maternal diabetes (MD) and maternal obesity (MO) have been robustly established to confer health risks in offspring. Additionally, mounting evidence suggests that these fetal programming effects vary by sex, but whether these factors independently or interactively influence infant brain development remains unclear.

**Objectives:** To characterize interactions between MD, MO, and sex on offspring subcortical brain volumes.

**Design, setting and participants:** This was a cross-sectional study of 1,966 infants from six international cohorts.

**Exposures:** MD and MO

**Main outcomes and measures:** MRI-based subcortical brain volumes (thalamus, amygdala, hippocampus, pallidum, putamen, caudate) were segmented and mixed effects models were used to examine associations, controlling for age at scan, prematurity, birthweight, maternal education, and intracranial volume. Backward elimination regression was used to identify the best fitting model (3-way interaction, 2-way interaction, no interaction) for each region and false discovery rate (FDR) corrections were applied.

**Results:** Of 1,966 infants, 46% were female (N=909), 9% were exposed to MD (N=172), and 21% were exposed to MO (N=386). MRI scans were performed at (mean±SD) 25.9±18.8 days of age. There was a significant interaction between MD, MO and sex in the thalamus (standardized β=−0.32, 95%CI −0.54 to −0.11, FDR corrected *P*=0.014). In female infants, MD (standardized β=−0.10, 95%CI –0.02 to −0.003, *P*=0.04) and MO (standardized β =−0.09, 95%CI −0.14 to –0.03, *P*=0.003) were independently and negatively associated with thalamic volume. In males, a significant interaction between MD and MO was observed (standardized β =−0.20, 95%CI −0.34 to –0.06, *P*=0.005), with post hoc analysis showing that males with combined exposure to MD and MO had lower thalamic volume compared to those with one or neither exposure (all *Ps*<0.05). In the hippocampus, an interaction between MO and infant sex was identified (standardized β =0.15, 95%CI 0.05 to 0.26, FDR corrected *P*=0.015), whereby MO (independent of MD) was associated with lower offspring hippocampal volume in females only (standardized β =−0.12, 95%CI −0.2 to −0.05, *P*=0.002).

**Conclusion and relevance:** Our results suggest independent, interactive associations of intrauterine exposure to MD and MO with infant subcortical brain volumes, varying by sex. This has implications for future metabolic disorders, among other health risks.

**Summary:** This study aims to investigate how sex modulates the influence of intrauterine exposure to maternal diabetes (MD) and maternal obesity (MO) on infant subcortical brain volumes. We observed sex-specific associations of gestational exposure to MD or MO with infant brain volumes in regions critical for motivation, emotion, and signal integration. In female offspring, MD and MO were negatively and independently associated with thalamic volume, while MO was negatively associated with hippocampal volume. In males, combined exposure to MD and MO was associated with lower thalamic volume. Sex modulates the influence of prenatal exposure to MD and/or MO on early brain development. This has implications for future metabolic disorders, among other health risks.

## Introduction

The intrauterine environment plays a critical role in fetal brain development, which begins around the second gestational week and continues to birth. Several conditions during pregnancy including obesity and diabetes have been shown to confer increased health risks in offspring^1–4^ but their neurological contributions have not been robustly established.

Preliminary studies suggest an influence of maternal diabetes (MD) exposure on the brain. In rats, offspring born to mothers with diabetes show morphometric malformations of the hypothalamus^5^ while in humans, exposure to MD prenatally is associated with hypothalamic gliosis^6,7^, smaller subcortical structures (e.g., thalamus, caudate, putamen), lower cortical thickness (CT)^8^ and smaller cortical gray matter (GM) volume during late childhood^9^. However, the existing studies have limitations. First, studies in humans have been limited to *later periods of life* (ages 7 – 11 years) making it difficult to disentangle the contributing effects of prenatal exposures from postnatal environmental factors ^6–8,10–13^. Second, maternal obesity (MO) in pregnancy has also been associated with alterations in offspring brains at birth and during infancy^14–20^ but prior studies have not disentangled the effects of MO from those driven independently by MD ^7,8,13,21–23^ and those resulting from combined diabetes and obesity exposure. Furthermore, there is a strong rationale for investigating sex-specific effects in the context of MO/MD as fetal development constitutes a critical period^24^ during which steroid hormone exposure drives sex-specific maternal placental function^25^, and epigenetic modification^26^. Indeed, these mechanisms underpin observed sex-specific differences in offspring brain outcomes including rodent models demonstrating MD/MO-related sex-specific gene expression changes^27–29^ in the brain, and human studies identifying sex-specific associations between MO and offspring hippocampal volume during childhood^30^. Yet, most human studies to date ^6,10–12,21–23^ have used *small sample sizes* and *homogeneous populations*, calling into question the reproducibility and generalizability of findings.

To address these knowledge gaps, we leveraged mega-analytics using a large international cohort to investigate associations between prenatal exposure to MD and/or MO and infant subcortical volumes, testing for moderating effects of sex. Based on prior studies^8,11,13^, we hypothesized that prenatal exposure to MD or MO would be associated with smaller subcortical region volumes, and combined exposure would exacerbate these effects. We also hypothesized that these associations may vary in a sex-dependent manner.

## Results

A total of 1,966 participants from 6 international cohorts were included (**Table 1**). Around 46% of the participants were female, 9% of infants were exposed to MD, and 22% were exposed to MO, prenatally. MRI scans were performed between 1 and 99 days old (mean = 25.87; SD = 18.77). The age distribution is shown in eFigure 1.

**Table 1:** Socio-demographic and birth outcome distribution of participant.

| Cohort |  | All | Cape Town | dHCP | FinnBrain | GMIA | UC Irvine | EBDS |
| --- | --- | --- | --- | --- | --- | --- | --- | --- |
| N |  | 1966 | 139 | 605 | 125 | 106 | 83 | 907 |
| Sex | Male | 1056 | 71 | 322 | 70 | 55 | 49 | 489 |
|  | Female | 909 | 68 | 283 | 55 | 51 | 34 | 418 |
| Diabetes Exposure | Yes | 172 | 1 | 49 | 16 | 5 | 6 | 95 |
|  | No | 1790 | 138 | 556 | 107 | 101 | 77 | 811 |
| Obesity Exposure | Yes | 386 | 51 | 74 | 13 | 10 | 21 | 217 |
|  | No | 1447 | 86 | 531 | 110 | 95 | 62 | 563 |
| Gestation | Preterm | 492 | 12 | 137 | 0 | 0 | 6 | 337 |
|  | Full-term | 1472 | 127 | 468 | 125 | 106 | 76 | 570 |
| Birthweight | Low | 427 | 6 | 136 | 0 | 0 | 4 | 281 |
|  | Normal | 1537 | 133 | 469 | 125 | 106 | 79 | 625 |
| Maternal Education | Primary-Secondary | 580 | 132 | 34 | 72 | 13 | 56 | 302 |
|  | Tertiary | 1359 | 7 | 549 | 49 | 93 | 27 | 605 |
| Family Income | Low | 518 | 47 | NA | 44 | 28 | 45 | 354 |
|  | Medium-High | 761 | 92 |  | 77 | 78 | 34 | 480 |

### ICV

MO was negatively associated with ICV in offspring, independent of MD (standardized β =−0.12, 95% CI −0.19 to −0.05, FDR corrected *P* =0.007). The association between MD and offspring ICV was non-significant after controlling for MO (standardized β =−0.06, 95% CI −0.16 to 0.04, FDR corrected *P* =0.261) (extended table 2).

**Table 2:** Regression results from the models selected for ICV, Hippocampus, and Thalamus.

|  | ICV |  |  |  |
| --- | --- | --- | --- | --- |
|  | Standardized Estimate | 95%CI (Standardized) | P-value | FDR-Adjusted P-value |
| Sex (Male) | 0.31 | 0.26 to 0.37 | **<0.0001** | **<0.0001** |
| MO | -0.12 | -0.19 to -0.05 | **0.0008** | **0.007** |
| MD | -0.06 | -0.16 to 0.04 | 0.261 | 0.386 |
| N | 1724 |  |  |  |
|  | Hippocampus |  |  |  |
|  | Standardized Estimate | 95%CI (Standardized) | P-value | FDR-Adjusted P-value |
| Sex (Male) | -0.01 | -0.06 to 0.04 | 0.763 | 0.858 |
| MO | -0.13 | -0.20 to -0.05 | 0.001 | 0.007 |
| MD | -0.05 | -0.13 to 0.02 | 0.170 | 0.321 |
| Sex (Male): MO | 0.15 | 0.05 to 0.26 | 0.004 | 0.015 |
| N | 1706 |  |  |  |
|  | Thalamus |  |  |  |
|  | Standardized Estimate | 95%CI (Standardized) | P-value | FDR-Adjusted P-value |
| Sex (Male) | -0.06 | -0.09 to -0.03 | 0.0004 | 0.004 |
| MO | -0.09 | -0.14 to -0.04 | 0.001 | 0.007 |
| MD | -0.08 | -0.17 to 0.01 | 0.089 | 0.218 |
| MO: MD | 0.13 | -0.03 to 0.29 | 0.104 | 0.236 |
| Sex (Male) : MO | 0.05 | -0.02 to 0.13 | 0.167 | 0.321 |
| Sex (Male) : MD | 0.11 | -0.02 to 0.24 | 0.083 | 0.218 |
| Sex (Male) : MO : MD | -0.32 | -0.54 to -0.11 | 0.003 | 0.014 |
| N | 1657 |  |  |  |

### Thalamus

Sex-specific associations of prenatal exposure to MD and MO on offspring thalamus were observed (sex-by-MO-by-MD interaction: standardized β =−0.32, 95% CI −0.54 to −0.11, FDR corrected *P* =0.014, **Figure 1**, **Table 2, eTable2**). In male infants, a synergistic MOxMD interaction in the thalamus was identified (standardized β =−0.20, 95% CI −0.34 to −0.06, *P* = 0.005), with post hoc analysis revealing that male infants with MO and MD had significantly lower thalamus volumes than male infants with either MD (standardized β = 0.89, 95% CI 0.31 to 1.46, *P* = 0.002) or MO alone (standardized β = 0.56, 95% CI 0.08 to 1.04, *P* = 0.031) or male infants exposed to neither (standardized β = 0.72, 95% CI 0.27 to 1.17, *P* = 0.719) (eTable 3). MD alone (standardized β = −0.17, 95% CI −0.54 to 0.21, *P* = 0.719) and MO alone (standardized β = 0.16, 95% CI −0.06 to 0.38, *P* = 0.328) were not significantly different from the un-exposed group (eTable 2). In female infants, MO (standardized β=−0.09, 95% CI −0.14 to −0.03, *P*=0.003) and MD (standardized β=−0.1, 95% CI −0.2 to −003, *P*=0.04) were negatively and independently associated with thalamic volume (eTable 2).

**Figure 1:**
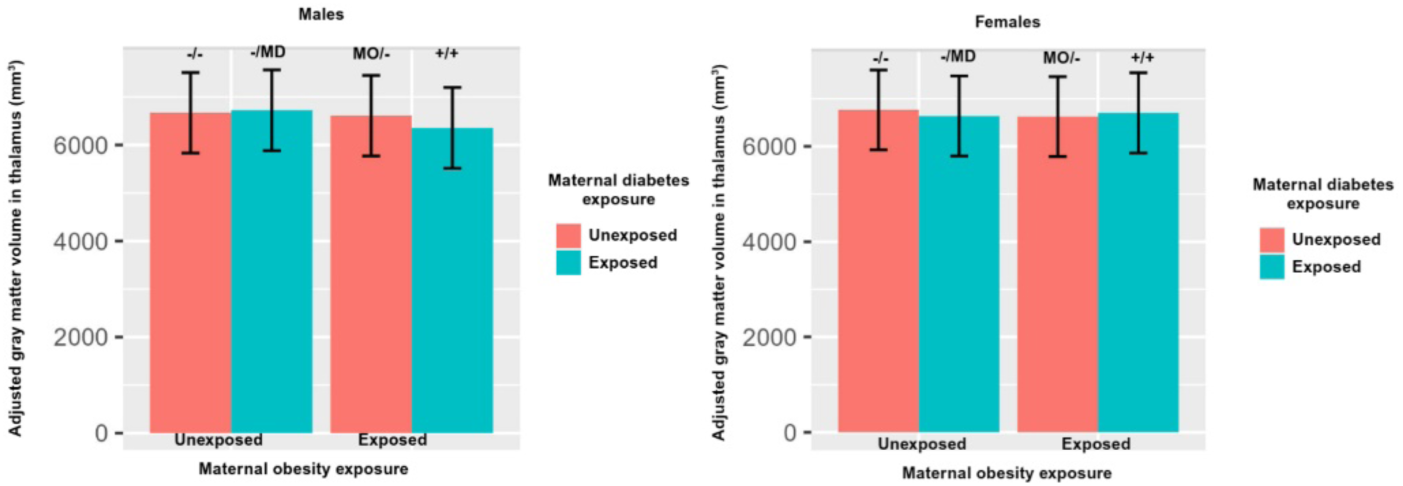
Pairwise comparisons of thalamic volume in male and female children exposed/unexposed to maternal obesity/diabetes. −/− denotes unexposed to maternal obesity or maternal diabetes; −/MD denotes exposed to maternal diabetes and not obesity; MO/− denotes exposed to maternal obesity and not diabetes; +/+ denotes exposed to both maternal diabetes and obesity. The number of observations in each category is as follows. Males: −/− (N=664), −/MD (N=43), MO/− (N=153), +/+ (N=30). Females: −/− (N=557), −/MD (N=40), MO/− (N=146), +/+ (N=24). The bar represents mean±SE in each group.

### Hippocampus

There was a significant sex-by-MO interaction on hippocampal volume (standardized β =0.15, 95% CI 0.05 to 0.26, FDR corrected *P* = 0.015) (**Table 2**, **Figure 2**). Sex-stratified analysis revealed an association between MO and hippocampal volume, independent of MD (standardized β =−0.12, 95% CI −0.20 to −0.05, *P* = 0.002) (eTable 4). Post-hoc analysis revealed that female infants with MO had significantly lower hippocampal volume than females not exposed to MO (standardized β =0.29, 95% CI 0.08 to 0.5, *P* = 0.007) (eTable 5), regardless of MD exposure.

**Figure 2:**
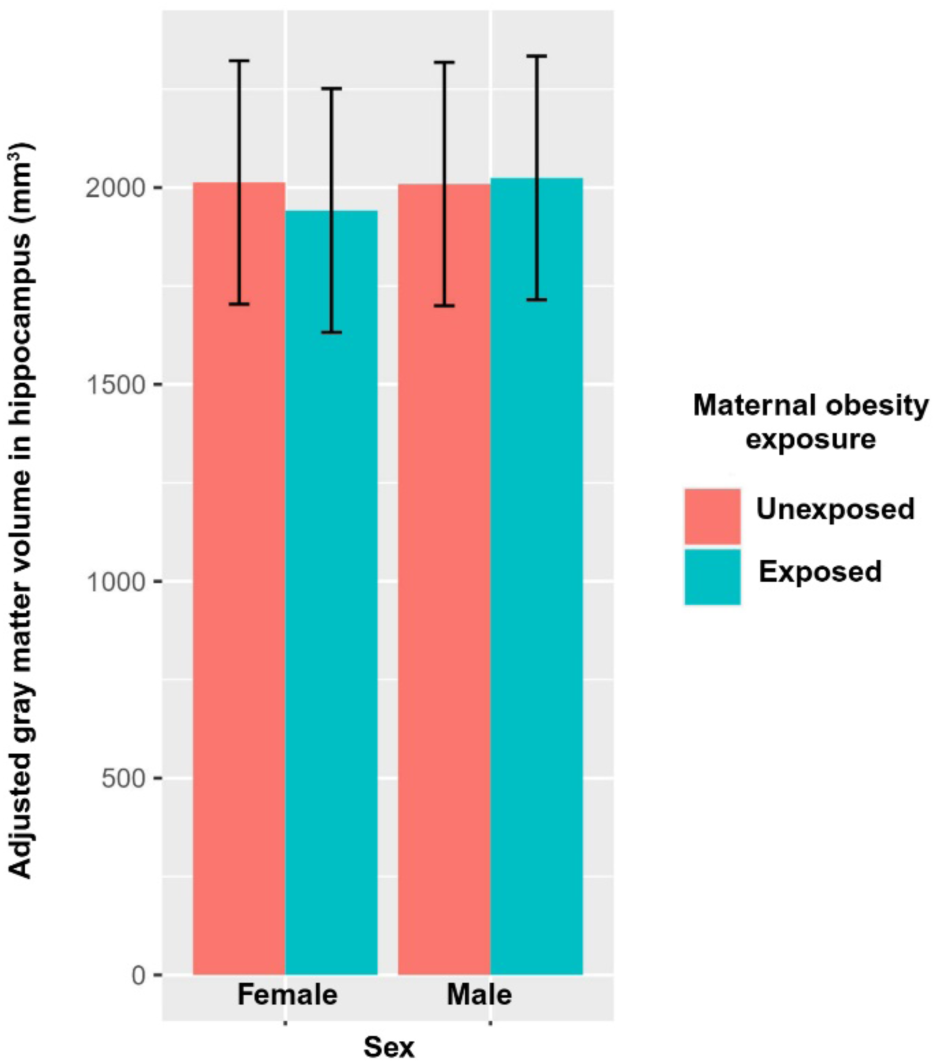
Pairwise comparisons of hippocampal volume in male and female children exposed/unexposed to maternal obesity. The number of observations in each category is as follows. Female-Obesity unexposed (N=624), Female –Obesity exposed (N=170), Male – Obesity unexposed (N=728), and Male – Obesity exposed (N=184). The bar represents mean±SE in each group.

We did not observe any associations between MD, MO and volumes of other subcortical regions (Extended Table 2). To allow for comparisons with prior studies, we present results in eTable 6 where models included MO only, MD only and MO+MD.

### Sensitivity analysis results

Excluding ICV (eTable 7) showed similar results for the thalamus and hippocampus. Amygdala (standardized β =−0.06, 95% CI −0.11 to −0.01, FDR corrected *P* = 0.027) and caudate (standardized β =−0.11, 95% CI −0.19 to −0.04, FDR corrected *P* = 0.01) volumes were negatively associated with MO, independent from MD, suggesting these associations may be dependent on ICV, in other words, non-specific to the amygdala and caudate. Excluding birthweight from the model did not alter outcomes in the ICV, thalamus and hippocampus (eTable 8). In the sensitivity analysis with only full-term infants, similar associations were observed for hippocampus and thalamus (eTable 9). Exclusion of DCHS cohort also showed similar data patterns for ICV and thalamus (eTable 10). Cohort-wise analysis exhibited consistent patterns across cohorts (eFigure 2).

## Discussion

In this large, well-powered, multi-cohort study we identified independent, interactive sex-dependent associations between *in utero* exposure to MD and/or MO and thalamic and hippocampal volumes. To our knowledge, this is the largest study to establish associations between prenatal exposure to MD and/or MO with subcortical brain volumes in early infancy. While ours is not the first study to report additive effects driven by MD and MO exposure, ^44,45^ it is the first to robustly establish this link with offspring brain development in early infancy.

MD and MO are the most common medical conditions during pregnancy and have separately been associated with structural and functional alterations in offspring brain during gestation and infancy ^14–20,22,23^. However, prior studies have failed to account for their frequent co-occurrence in exploring relationships with offspring brain outcomes. Our study marks an important advancement, explicitly recognizing this critical confound and disentangling the influence of MD from that of MO. Interestingly, the most prominent findings of our study are sex specific associations of MD and MO with offspring thalamic and hippocampal volumes. In females, MD and MO were independently and negatively related to thalamus volumes, but combined exposures were not. In contrast, in males we observed that combined exposure was associated with lower thalamic volumes with moderate to large effect sizes, but MD or MO alone were not. We also found a significant interaction between hippocampal volumes, MO exposure, and sex, which was driven by lower volumes in female offspring exposed to MO. These findings are noteworthy as sex differences in the thalamus and hippocampus could provide the biological basis for studies that have reported sex differences in cardiometabolic disorders and neurodevelopmental disorders following prenatal exposure to MD and MO^46^. While the current study was not designed to identify the biological mechanisms underlying these sex-specific effects, there is a rich body of evidence demonstrating that prenatal steroid hormone exposure drives sex-specific maternal placental function^25^ and epigenetic modification^26^ in the developing brain. It then follows that the sex-specific findings reported here may result from an interaction between upstream hormonally programmed sex differences in the brain and exposure to diabetes/obesity-related ligands. Further studies would be needed to explore this possibility. Regardless, the current findings illustrate the important regulatory role of sex in offspring brain outcomes and emphasize the need to factor sex into analyses exploring these metrics.

According to Developmental Origins of Health and Disease theory^47^, environmental exposures act in the earliest stage of life to influence health and disease later in life and the developing brain has been shown to be a target of these exposures. In our study, the thalamus and hippocampus show the greatest changes under the influence of MD or MO. Thalamic development starts as early as 8-14 weeks of gestation and has the most protracted period of development among subcortical regions^36^, making it highly susceptible to environmental insults. Moreover, our results are in concordance with a recent study in which 9-10-year-old children with prenatal exposure to MD had smaller thalamic volumes than un-exposed children ^9^. However, that study did not account for MO exposure, nor did it examine interactions with sex. Our results extend this work and provide a more robust scaffold for our understanding of this developmental trajectory. The hippocampus has also been reported to be susceptible to environmental insults in infant imaging studies ^47–51^ but the findings have been more mixed and there is thus a lack of consensus. In contrast with our work, a study in 7-11-year-old children exposed to MO prenatally showed that males but not females had lower hippocampal volumes^30^, but this study was limited by sample size and studied older children, whereas our study included one of the largest samples of infant imaging near birth without confounding contributors of postnatal exposures. In a separate study, lower hippocampal thickness was observed in 7–11-year-old children exposed to MD prenatally^11^, while three other studies in children with similar age ranges did not observe such associations ^9,11,13^. Our study aligns with the latter, confirming the absence of prenatal influence of MD exposure on hippocampal volume. Moreover, although a prior study in 9-10-year-old children reported significant negative relationships between prenatal exposure to MD and dorsal striatum volumes^9^ (i.e., putamen, caudate), we did not find such relationships in our younger cohort. We speculate that prior significant findings may be diminished after controlling for MO exposure given our data demonstrating that relationships between MD exposure and infant subcortical volumes were attenuated after adjusting for MO exposure. Furthermore, our sensitivity analyses emphasize the robustness of our findings.

Our study has strengths and limitations. First, a key strength is that we investigated relationships between prenatal exposure to MD and/or MO and the developing brain in early infancy, thereby reducing confounding postnatal factors. Second, as diabetes and obesity often go together during pregnancy, we studied both independent and combined associations of MD and MO with subcortical volumes. Third, we leveraged data from six international cohorts, providing the statistical power necessary to demonstrate reproducibility and estimates of effect sizes in the context of MD/MO in heterogeneous populations. Finally, we ruled out possible cohort effects by analyzing each cohort separately and observing the direction of effects--in turn demonstrating the robustness of results. Nonetheless, our study is limited by the self-reported status of MD and MO during pregnancy in some cohorts, lack of classification of types of diabetes during pregnancy, lack of information on maternal medication use, and lack of metabolic markers in offspring. The sample was over-represented for pre-term infants, and we cannot exclude other confounders such as maternal mental health in pregnancy which has also been associated with infant outcomes. We also restricted our analysis to subcortical volumes. Investigating other neuroimaging phenotypes, including thickness and surface area of cortical regions is an important future direction.

## Conclusions

In a diverse sample of 1,966 infants, prenatal exposure to maternal diabetes or maternal obesity was independently associated with lower thalamus volumes in females; Males with concomitant maternal diabetes and maternal obesity exposure demonstrated the lowest thalamic volumes. Females exposed to maternal obesity, but not males, exhibited lower hippocampus volumes. These results support the hypothesis that gestational exposure to maternal diabetes and/or maternal obesity influences the developing brain, and sex moderates these effects.

## STAR Methods

### Participant characteristics

The ENIGMA-ORIGINs working group^35,36^ contributed data from five cohorts: the Drakenstein Child Health Study (South Africa), UNC Early Brain Development Study (EBDS), UNC Gut Microbiome and Infant Anxiety (GMIA), the University of California, Irvine, USA, and the developing Human Connectome Project (UK). Additional data was provided by FinnBrain (Finland). Approval was obtained from respective local ethical review boards and informed consent from parents/legal guardians were obtained before data collection.

### Cohort characteristics

#### Drakenstein Child Health Study (DCHS)

This cohort was established to investigate the determinants of child growth, health, and development in a stable, semi-urban, low socioeconomic status community in South Africa. Exclusion criteria were minimal and focused on unavailability for follow-up. Detailed information is available elsewhere^31^.

#### UNC Early Brain Development Study (EBDS)

The cohort included an exceptionally large sample of typically developing infants, children at risk for schizophrenia and bipolar illness, a “structural” high risk group (children with prenatal isolated mild ventriculomegaly), and a sample of twins. Major medical illness in the mother, abnormality on ultrasound, and current substance abuse were exclusion criteria at enrollment. Detailed information is available elsewhere^32–34^.

#### UNC Gut Microbiome and Infant Anxiety (GMIA)

This is a prospective, longitudinal study of the microbiome-gut-brain axis. Recruitment was restricted to infants without major dietary or medical interventions. Inclusion criteria were infants born at term, vaginally delivered, and exclusively breastfed for the first 2 weeks. Exclusion criteria were admission to neonatal intensive care unit, weighing <2500 grams at birth, receiving antibiotics in the first 2 weeks, or metal in the body. Infants were excluded if the mother received antibiotics in the two weeks before delivery. Detailed information is available elsewhere^35^.

#### University of California, Irvine

This is a prospective, longitudinal, follow-up study in a population-based cohort. Exclusion criteria included preterm birth (< 34 completed weeks gestation), psychotropic medication, corticosteroids, smoking, and drug use during pregnancy (self-reports verified by urinary cotinine and drug toxicology), congenital or genetic disorders, and major neurologic disorder at birth. Detailed information is available elsewhere^36^.

#### Developing Human Connectome Project (dHCP)

This project aims to study neurodevelopment in a typical urban population sample from 20 to 44 weeks post-conception. Exclusion criteria include mothers or infants with contraindications to MR imaging, preterm infants too unwell to undergo scanning, and language barriers to obtaining consent. Detailed information is available elsewhere^37^.

#### FinnBrain

This is a birth cohort study recruiting Mother-infant dyads in Finland. Inclusion criteria were gestational age at birth ≥35 weeks and birth weight > 1500 g. Previously diagnosed central nervous system anomalies or abnormal findings in a previous MRI scan were exclusion criteria. Detailed information is available elsewhere^38,39^.

### Imaging Measures

Each site acquired and processed structural 3D T1-weighted and T2-weighted volumetric brain MRI scans of participants. Images were acquired at a field strength of 3T. Quality control was performed locally. Acquisition parameters and segmentation protocol details are provided in eTable 1.

### Exposure

Information on MD exposure during pregnancy including gestational diabetes mellitus, pre-gestational type 1 diabetes, and pre-gestational type 2 diabetes was either extracted from electronic medical records (EMR; in the EBDS, UC Irvine, dHCP, and FinnBrain cohorts) and/or self-reported. Only the UC Irvine and FinnBrain cohorts distinguished the type of MD. Pre-pregnancy BMI ≥ 30 kg/m^2^ was used to classify maternal obesity, and < 30 kg/m^2^ classified as non-obese.

### Statistical Analysis

To investigate the effects of MD and MO exposure on infant brain volumes, we used a sequence of mixed effects models that included the random effect of the cohort plus fixed effects of various exposures and covariates and their interactions. Our baseline model included the random effect of the cohort and fixed effects of MD and MO, sex, age at scan, gestational length (full-term vs pre-term birth), birthweight (normal vs low birthweight), and maternal education (tertiary education vs primary-secondary education). Additionally, for subcortical volumes we included ICV as a covariate.

To investigate modulating effects of sex, MO, and MD, we expanded the baseline model by adding first order (sex-by-MO, sex-by-MD, and MO-by-MD) and second order interactions (sex-by-MO-by MD). Thus, the full model included all main effects of the baseline model plus the first and second order interactions described above. Subsequently, using the full model, we performed backward elimination on interactions terms, first removing second order interactions if these interactions were not significant (1DF Likelihood Ratio Test, LRT, *p*-value<0.05). Then, if second order interactions were not significant, we eliminated 1st order interactions that had 1DF-LRT *p*-value<0.05. Final models included all the effects of the baseline model plus interactions that were retained after backward elimination. In the final models, if interactions with sex associations were significant, we followed up with sex-stratified analysis. Pairwise comparisons were performed using least-squares means method (lsmeans) and emmeans package^40^ in R. For effect size comparison we used Cohen’s d statistics^41^. All models were fitted using the lmer function of the lme4 R-package^42^ via maximum likelihood, and back-ward elimination was done using likelihood ratio tests using the ANOVA function of the baseline R-package.

To account for multiple testing, in final models we adjusted p-values using the Benjamini-Hochberg FDR correction^43^. We performed residual diagnostics to assess potential violations of models’ assumptions and found no evidence of strong departures from normality.

We performed sensitivity analyses to check the robustness of results. Sensitivity analysis 1: During the first 100 days, when the brain is developing at an exponential rate, ICV and subcortical structures may have unique growth patterns and adjusting for ICV may have different effects on each regional volume. To address this, we performed a sensitivity analysis by removing ICV from the models for subcortical volumes. Sensitivity analysis 2: We removed birthweight from the main model because birthweight is correlated with gestational term. Sensitivity analysis 3: Since preterm children constitute around a quarter of the sample and MO and MD are known risk factors for preterm birth, we performed a subsample analysis that included only full-term children. Sensitivity analysis 4: The DCHS cohort did not specifically have pre-pregnancy BMI, but rather maternal BMI. To investigate if effects of exposure to pre-pregnancy BMI were similar to exposure to maternal BMI, we performed analyses excluding the DCHS cohort from the dataset. Sensitivity analysis 5: We conducted a cohort-wise analysis to assess replicability of results.

## Data Availability

Six different cohorts contributed data for this project. Three cohorts have deposited data in the NIMH Data Archive (NDA) and investigators can gain access by submitting a Data Access Request to the NDA. Imaging data for twins in the EBDS cohort is available through NDA #1974 and NDA #2384 and for singletons via NDA #4314. Imaging data for UCI is available via NDA #1890. Data for dHCP is available via NDA#3955. Data from the other cohorts may be available upon request to the parent cohort contingent on getting appropriate IRB approval or data use agreements. Further information on ORIGINs dataset access and sharing can be obtained upon contact with the principal investigator (PI) Rebecca Knickmeyer.

## Acknowledgements

We thank all the participants and the families who contributed to this study. ORIGINs is supported by NIMH (R01MH123716 to RK) and the ENIGMA consortium is supported by the National Institutes of Health (NIH) Big Data to Knowledge (BD2K) award for foundational support and consortium development (Grant No. U54 EB020403 to PMT). This project is supported by NIDDK (R01DK137899 to SL). The independent cohorts within this study were supported by the grants and fellowships listed. DCHS - Bill & Melinda Gates Foundation (OPP 1017641), Medical Research Council South Africa: HJZ;Academy of Medical Sciences Newton Advanced Fellowship (NAF002/1001):KAD, the UK Government’s Newton Fund, by NIAAA via (R21AA023887), by the Collaborative Initiative on Fetal Alcohol Spectrum Disorders (CIFASD) developmental grant (U24 AA014811), US Brain and Behavior Foundation Independent Investigator grant (24467), and the Carnegie Corporation of New York, South African Medical Research Council (SA-MRC) – KAD, DJS, and HZ. EBDS – National Institutes of Health MH070890, HD053000, MH064065: JHG, and P50 HD103573: MS; UCI - R00 HD100593 (PI, Rasmussen); R01 MH091351 (PI, Buss); R01HD060628 (PI, Wadhwa). GMIA – National Institutes of Health R21 MH104330 and R33 MH104330: RCK. dHCP - European Research Council under the European Union Seventh Framework Programme (FP/20072013)/ERC Grant Agreement no. 319456. FinnBrain - Research Council of Finland (#325292); Brain and Behavior Research Foundation; NARSAD YI Grant (#1956); Finnish State Grants for Clinical Research; Signe and Ane Gyllenberg Foundation: LK; Research Council of Finland (#134950, #253270), Finnish State Grants for Clinical Research; Signe and Ane Gyllenberg Foundation, Jane and Aatos Erkko FoundationSigne and Ane Gyllenberg Foundation: HK; Sigrid Jusélius Foundation; Emil Aaltonen Foundation; Finnish Medical Foundation; Hospital District of Southwest Finland; State Grants for Clinical Research (ERVA); Signe and Ane Gyllenberg Foundation: JJT. KL is supported by K01MH135160.

## Author Contributions

SL conceptualized and obtained funding for this study; AA, GC, RK and SL performed statistical analysis; AA, JR, GC, RK and SL drafted original manuscript, and the rest of co-authors reviewed and edited it.

## Resource Availability Lead Contact

Further information and requests for resources and reagents should be directed to and will be fulfilled by the lead contact, Shan Luo.

## Materials Availability

This study did not generate new unique reagents.

## References

1. Kankowski L, Ardissino M, McCracken C, et al. The Impact of Maternal Obesity on Offspring Cardiovascular Health: A Systematic Literature Review. Front Endocrinol (Lausanne). 2022;13. doi:10.3389/FENDO.2022.868441

2. Robles MC, Campoy C, Fernandez LG, Lopez-Pedrosa JM, Rueda R, Martin MJ. Maternal Diabetes and Cognitive Performance in the Offspring: A Systematic Review and Meta-Analysis. PLoS One. 2015;10(11). doi:10.1371/JOURNAL.PONE.0142583

3. Kong L, Chen X, Gissler M, Lavebratt C. Relationship of prenatal maternal obesity and diabetes to offspring neurodevelopmental and psychiatric disorders: a narrative review. Int J Obes (Lond). 2020;44(10):1981–2000. doi:10.1038/S41366-020-0609-4

4. Bianco ME, Josefson JL. Hyperglycemia During Pregnancy and Long-Term Offspring Outcomes. Curr Diab Rep. 2019;19(12). doi:10.1007/S11892-019-1267-6

5. Plagemann A, Harder T, Janert U, et al. Malformations of hypothalamic nuclei in hyperinsulinemic offspring of rats with gestational diabetes. Dev Neurosci. 1999;21(1):58–67. doi:10.1159/000017367

6. Chandrasekaran S, Melhorn S, Olerich KLW, et al. Exposure to Gestational Diabetes Mellitus Prior to 26 Weeks Is Related to the Presence of Mediobasal Hypothalamic Gliosis in Children. Diabetes. 2022;71(12):2552–2556. doi:10.2337/DB22-0448

7. Olerich K, Sewaybricker L, Chandrasekaran S, Melhorn S, Kee S, Schur EA. Association of maternal diabetes with offspring childhood hypothalamic gliosis. Am J Obstet Gynecol. 2022;226(1):S157–S158. doi:10.1016/j.ajog.2021.11.276

8. Ahmed S, Cano MÁ, Sánchez M, Hu N, Ibañez G. Effect of exposure to maternal diabetes during pregnancy on offspring’s brain cortical thickness and neurocognitive functioning. Child Neuropsychol. 2023;29(4):588–606. doi:10.1080/09297049.2022.2103105

9. Nivins S, Klingberg T. Effects of prenatal exposure to maternal diabetes mellitus on deep grey matter structures and attention deficit hyperactivity disorder symptoms in children. Acta Paediatr. 2023;112(7):1511–1523. doi:10.1111/APA.16756

10. Page KA, Luo S, Wang X, et al. Children Exposed to Maternal Obesity or Gestational Diabetes Mellitus During Early Fetal Development Have Hypothalamic Alterations That Predict Future Weight Gain. Diabetes Care. 2019;42(8):1473–1480. doi:10.2337/DC18-2581

11. Lynch KM, Alves JM, Chow T, et al. Selective morphological and volumetric alterations in the hippocampus of children exposed in utero to gestational diabetes mellitus. Hum Brain Mapp. 2021;42(8):2583–2592. doi:10.1002/HBM.25390

12. Luo S, Angelo BC, Chow T, et al. Associations Between Exposure to Gestational Diabetes Mellitus In Utero and Daily Energy Intake, Brain Responses to Food Cues, and Adiposity in Children. Diabetes Care. 2021;44(5):1185–1193. doi:10.2337/DC20-3006

13. Luo S, Hsu E, Lawrence KE, et al. Associations among prenatal exposure to gestational diabetes mellitus, brain structure, and child adiposity markers. Obesity. 2023;31(11):2699–2708. doi:10.1002/OBY.23901

14. Ou X, Thakali KM, Shankar K, Andres A, Badger TM. Maternal Adiposity Negatively Influences Infant Brain White Matter Development. Obesity (Silver Spring). 2015;23(5):1047. doi:10.1002/OBY.21055

15. Verdejo-Román J, Björnholm L, Muetzel RL, et al. Maternal prepregnancy body mass index and offspring white matter microstructure: results from three birth cohorts. Int J Obes (Lond). 2019;43(10):1995–2006. doi:10.1038/S41366-018-0268-X

16. Li X, Andres A, Shankar K, et al. Differences in brain functional connectivity at resting state in neonates born to healthy obese or normal-weight mothers. Int J Obes (Lond). 2016;40(12):1931–1934. doi:10.1038/IJO.2016.166

17. Salzwedel AP, Gao W, Andres A, et al. Maternal Adiposity Influences Neonatal Brain Functional Connectivity. Front Hum Neurosci. 2019;12. doi:10.3389/FNHUM.2018.00514

18. Rajasilta O, Häkkinen S, Björnsdotter M, et al. Maternal pre-pregnancy BMI associates with neonate local and distal functional connectivity of the left superior frontal gyrus. Scientific Reports 2021 11:1. 2021;11(1):1–9. doi:10.1038/s41598-021-98574-9

19. Na X, Phelan NE, Tadros MR, et al. Maternal Obesity during Pregnancy is Associated with Lower Cortical Thickness in the Neonate Brain. AJNR Am J Neuroradiol. 2021;42(12):2238–2244. doi:10.3174/AJNR.A7316

20. Spann MN, Scheinost D, Feng T, et al. Association of Maternal Prepregnancy Body Mass Index With Fetal Growth and Neonatal Thalamic Brain Connectivity Among Adolescent and Young Women. JAMA Netw Open. 2020;3(11). doi:10.1001/JAMANETWORKOPEN.2020.24661

21. Linder K, Schleger F, Kiefer-Schmidt I, et al. Gestational Diabetes Impairs Human Fetal Postprandial Brain Activity. J Clin Endocrinol Metab. 2015;100(11):4029–4036. doi:10.1210/JC.2015-2692

22. Avci R, Whittington JR, Blossom SJ, et al. Studying the Effect of Maternal Pregestational Diabetes on Fetal Neurodevelopment using Magnetoencephalography. Clin EEG Neurosci. 2020;51(5):331. doi:10.1177/1550059420909658

23. Xuan DS, Zhao X, Liu YC, et al. Brain Development in Infants of Mothers With Gestational Diabetes Mellitus: A Diffusion Tensor Imaging Study. J Comput Assist Tomogr. 2020;44(6):947–952. doi:10.1097/RCT.0000000000001110

24. Bakker J. The role of steroid hormones in the sexual differentiation of the human brain. J Neuroendocrinol. 2022;34(2). doi:10.1111/JNE.13050

25. Bale TL. The placenta and neurodevelopment: sex differences in prenatal vulnerability. Dialogues Clin Neurosci. 2016;18(4):459–464. doi:10.31887/DCNS.2016.18.4/TBALE

26. McCarthy MM, Auger AP, Bale TL, et al. The epigenetics of sex differences in the brain. J Neurosci. 2009;29(41):12815–12823. doi:10.1523/JNEUROSCI.3331-09.2009

27. Edlow AG, Guedj F, Pennings JLA, Sverdlov D, Neri C, Bianchi DW. Males are from Mars, females are from Venus: sex-specific fetal brain gene expression signatures in a mouse model of maternal diet-induced obesity. Am J Obstet Gynecol. 2016;214(5):623.e1. doi:10.1016/J.AJOG.2016.02.054

28. Aviel-Shekler K, Hamshawi Y, Sirhan W, et al. Gestational diabetes induces behavioral and brain gene transcription dysregulation in adult offspring. Translational Psychiatry 2020 10:1. 2020;10(1):1–11. doi:10.1038/s41398-020-01096-7

29. Sousa FJ, Correia RG, Cruz AF, et al. Sex differences in offspring neurodevelopment, cognitive performance and microglia morphology associated with maternal diabetes: Putative targets for insulin therapy. Brain Behav Immun Health. 2020;5. doi:10.1016/J.BBIH.2020.100075

30. Alves JM, Luo S, Chow T, Herting M, Xiang AH, Page KA. Sex differences in the association between prenatal exposure to maternal obesity and hippocampal volume in children. Brain Behav. 2020;10(2). doi:10.1002/BRB3.1522

31. Donald KA, Wedderburn CJ, Barnett W, et al. Risk and protective factors for child development: An observational South African birth cohort. Batura N, ed. PLoS Med. 2019;16(9):e1002920. doi:10.1371/journal.pmed.1002920

32. Knickmeyer RC, Xia K, Lu Z, et al. Impact of Demographic and Obstetric Factors on Infant Brain Volumes: A Population Neuroscience Study. Cerebral Cortex. 2016;27(12):5616–5625. doi:10.1093/cercor/bhw331

33. Gilmore JH, Schmitt JE, Knickmeyer RC, et al. Genetic and environmental contributions to neonatal brain structure: A twin study. Hum Brain Mapp. 2010;31(8):1174–1182. doi:10.1002/hbm.20926

34. Knickmeyer RC, Gouttard S, Kang C, et al. A structural MRI study of human brain development from birth to 2 years. Journal of Neuroscience. 2008;28(47):12176–12182. doi:10.1523/JNEUROSCI.3479-08.2008

35. Carlson AL, Xia K, Azcarate-Peril MA, et al. Infant gut microbiome composition is associated with non-social fear behavior in a pilot study. Nat Commun. 2021;12(1). doi:10.1038/S41467-021-23281-Y

36. Alex AM, Aguate F, Botteron K, et al. A global multicohort study to map subcortical brain development and cognition in infancy and early childhood. Nature Neuroscience 2023 27:1. 2023;27(1):176–186. doi:10.1038/s41593-023-01501-6

37. Edwards AD, Rueckert D, Smith SM, et al. The Developing Human Connectome Project Neonatal Data Release. Front Neurosci. 2022;16. doi:10.3389/FNINS.2022.886772

38. FinnBrain study. Accessed November 20, 2024. https://sites.utu.fi

39. Karlsson L, Tolvanen M, Scheinin NM, et al. Cohort Profile: The FinnBrain Birth Cohort Study (FinnBrain). Int J Epidemiol. 2018;47(1):15–16j. doi:10.1093/IJE/DYX173

40. Lenth R V. emmeans: Estimated Marginal Means, aka Least-Squares Means. Published online 2024. Accessed November 14, 2024. https://rvlenth.github.io/emmeans/

41. Cohen J. Things I have learned (so far). American Psychologist. 1990;45(12):1304–1312. doi:10.1037/0003-066X.45.12.1304

42. Bates D, Mächler M, Bolker BM, Walker SC. Fitting Linear Mixed-Effects Models Using lme4. J Stat Softw. 2015;67(1):1–48. doi:10.18637/JSS.V067.I01

43. Benjamini Y, Hochberg Y. Controlling the False Discovery Rate: A Practical and Powerful Approach to Multiple Testing. J R Stat Soc Series B Stat Methodol. 1995;57(1):289–300. doi:10.1111/J.2517-6161.1995.TB02031.X

44. Pirkola J, Pouta A, Bloigu A, et al. Risks of Overweight and Abdominal Obesity at Age 16 Years Associated With Prenatal Exposures to Maternal Prepregnancy Overweight and Gestational Diabetes Mellitus. Diabetes Care. 2010;33(5):1115–1121. doi:10.2337/DC09-1871

45. Kong L, Nilsson IAK, Gissler M, Lavebratt C. Associations of Maternal Diabetes and Body Mass Index With Offspring Birth Weight and Prematurity. JAMA Pediatr. 2019;173(4):371–378. doi:10.1001/JAMAPEDIATRICS.2018.5541

46. Talbot CPJ, Dolinsky VW. Sex Differences in the Developmental Origins of Cardiometabolic Disease Following Exposure to Maternal Obesity and Gestational Diabetes. 2018. doi:10.1139/APNM-2018-0667

47. Gluckman PD, Hanson MA, Cooper C, Thornburg KL. Effect of in utero and early-life conditions on adult health and disease. N Engl J Med. 2008;359(1):61–73. doi:10.1056/NEJMRA0708473

48. Salzwedel AP, Grewen KM, Vachet C, Gerig G, Lin W, Gao W. Prenatal Drug Exposure Affects Neonatal Brain Functional Connectivity. Journal of Neuroscience. 2015;35(14):5860–5869. doi:10.1523/JNEUROSCI.4333-14.2015

49. Rotem-Kohavi N, Williams LJ, Muller AM, et al. Hub distribution of the brain functional networks of newborns prenatally exposed to maternal depression and SSRI antidepressants. Depress Anxiety. 2019;36(8):753–765. doi:10.1002/DA.22906

50. Scheinost D, Spann MN, McDonough L, Peterson BS, Monk C. Associations between different dimensions of prenatal distress, neonatal hippocampal connectivity, and infant memory. Neuropsychopharmacology. 2020;45(8):1272–1279. doi:10.1038/S41386-020-0677-0

51. Sheridan MA, How J, Araujo M, Schamberg MA, Nelson CA. What are the links between maternal social status, hippocampal function, and HPA axis function in children? Dev Sci. 2013;16(5):665–675. doi:10.1111/DESC.12087

