## Supplementary Figures for "Infant Subcortical Brain Volumes Associated with Maternal Obesity and Diabetes: A Large Multicohort Study"

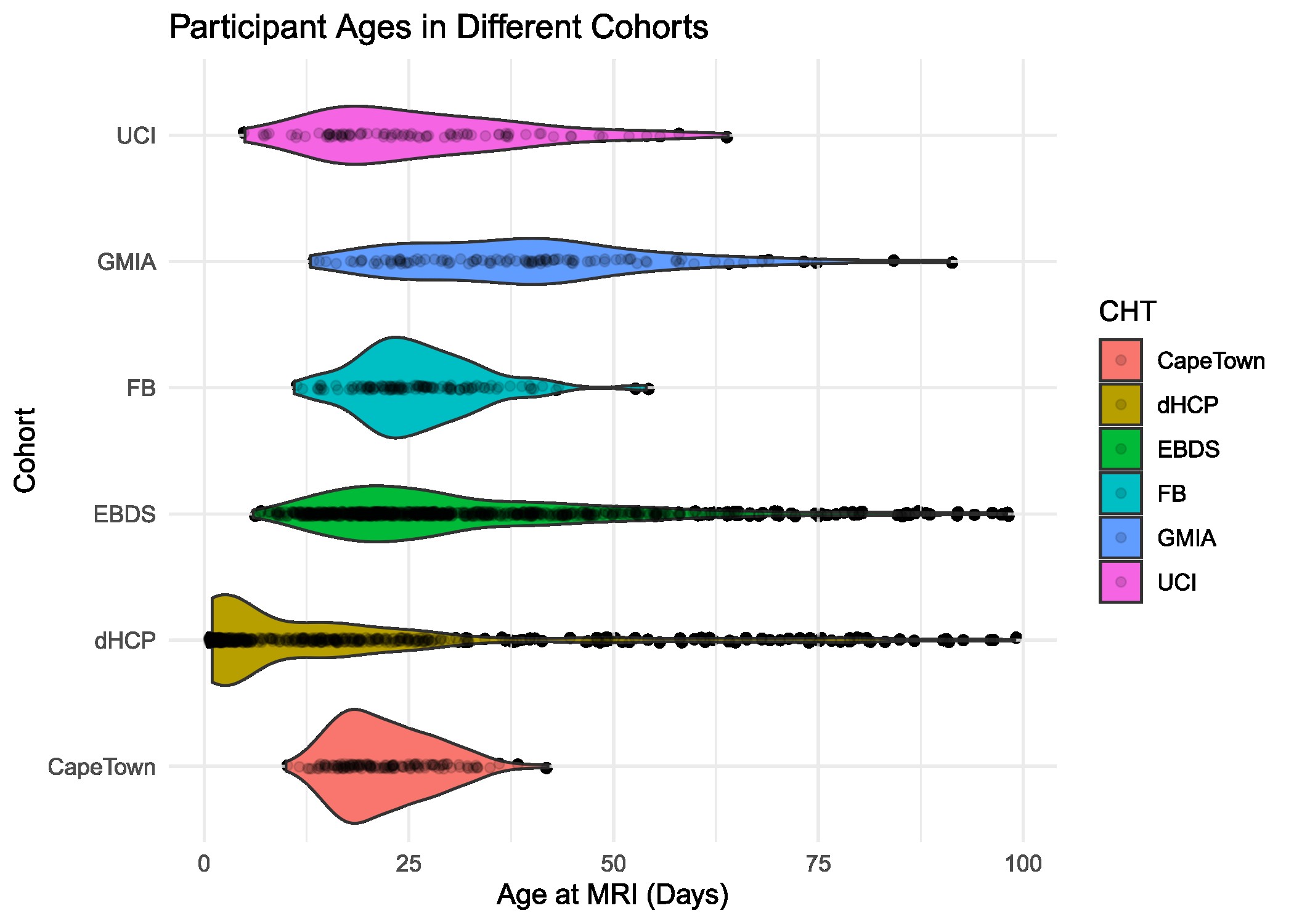
eFigure1: Distribution of age at scan for the participants from the different cohorts. Number of individuals for UCI is 83, GMIA is 106, FB (FinnBrain) is 125, EBDS is 907, dHCP is 605, and Cape Town (DCHS) is 139.

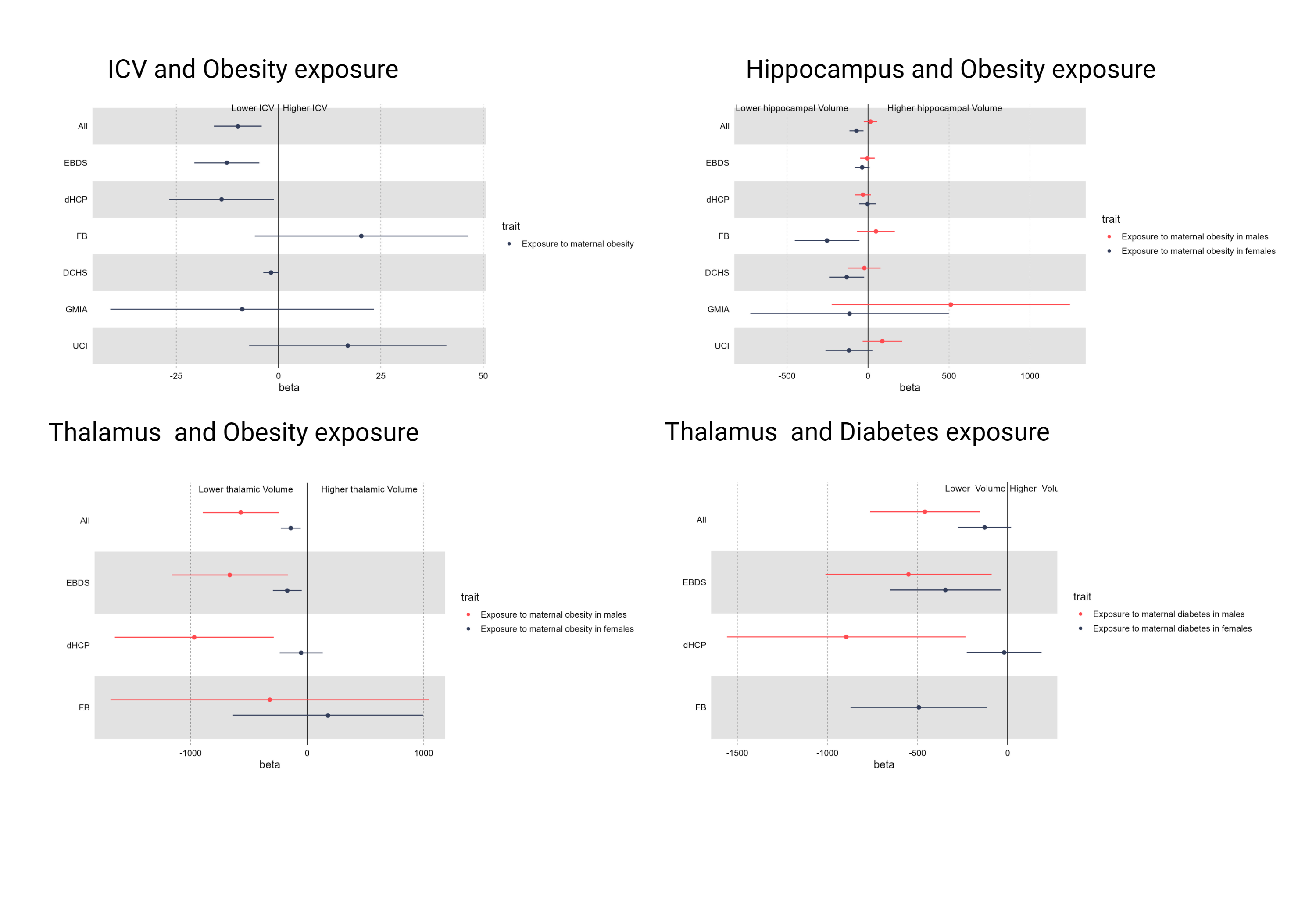
eFigure 2: Forest plot illustrating the association of MO and MD with ICV, hippocampus and thalamus across the different cohorts. The estimates (beta values) are plotted with 95% confidence intervals represented by the horizontal lines. The illustration is stratified by trait. Negative beta values represent association with lower volume and positive beta values represent association with higher volumes. The Red color corresponds to associations in males and black represent associations in females.
